# Unexpected Increase in Postoperative Atrial Fibrillation by Calcium-mediated Autonomic Denervation: Results of the CAP-AF2 Trial

**DOI:** 10.1101/2023.06.05.23290999

**Authors:** Huishan Wang, Yuji Zhang, Fangran Xin, Jikai Zhao, Keyan Zhao, Dengshun Tao, Praloy Chakraborty, Zongtao Yin, Guannan Liu, Sunny S. Po

## Abstract

**Background:** In the CAP-AF trial, injection of calcium chloride (CaCl_2_) into the four major atrial ganglionated plexi (GP) reduced the relative risk of postoperative atrial fibrillation (POAF) by 63% in patients undergoing coronary artery bypass surgery (CABG).

**Objective:** The CAP-AF2 trial intended to investigate if similar autonomic denervation could prevent POAF in patients with mitral regurgitation (MR) but without persistent AF who underwent surgery for MR.

**Methods:** The CAP-AF-2 trial was an investigator-initiated, single center, double-blind, randomized clinical trial. This trial planned to 1:1 randomize 320 adult patients to CaCl_2_ vs. sodium chloride (NaCl, sham) injection into the four GP during surgery. The primary outcome was incidence of POAF (≥30 seconds) in 7 days. Secondary outcomes included length of hospitalization, POAF burden, actionable antiarrhythmic therapy for POAF, heart rate variability changes and plasma inflammatory markers.

**Results:** This trial was terminated after midterm analysis showing that the cumulative POAF incidence was higher in the CaCl_2_ group (43/78, 55.13%) than the NaCl group (31/82, 37.80%; confidence interval of difference 1.01%-32.48%, *P*= 0.028). In the CaCl_2_ group, more patients were hospitalized over 7 days (69.8% vs. 45.2%; p=0.033) and more patients required amiodarone therapy (p=0.039). AF burden, plasma inflammatory markers and heart rate variability were not different between the two groups. Frequent atrial bigeminy or nonsustained atrial tachycardia immediately preceded POAF in 76.7% (CaCl_2_) and 29.0% (NaCl) patients, respectively (*P*<0.001), consistent with triggers caused by higher sympathetic activity. Immunohistochemistry study obtained from GP and left atrium during surgery revealed parasympathetic dominance in patients receiving MV surgery but sympathetic dominance in patients undergoing CABG.

**Conclusions:** Unlike patients undergoing CABG, autonomic denervation increased the risk of POAF in patients receiving MR surgery, indicating distinct AF substrate in different cardiovascular diseases. Calcium-mediated autonomic denervation may have tipped the tissue autonomic balance toward sympathetic dominance and provided triggers for POAF. While autonomic denervation has emerged as a novel therapy to treat various cardiovascular diseases, it should only be performed with evidence supported by randomized clinical trials.

**The Chinese Clinical Trial Registry registration number:** ChiCTR2000029314.

**website**: http://www.chictr.org.cn/showproj.aspx?proj=48587

**CLINICAL PERSPECTIVES:** *What is new:*

- Calcium-mediated autonomic denervation increased the incidence of post-operative atrial fibrillation (AF) in patient undergoing mitral valve surgery for severe mitral regurgitation, contradictory to the beneficial effects it exerted on patients undergoing coronary artery bypass surgery.

*Clinical implications:*

- Each cardiovascular disease may have its distinct autonomic balance at the tissue level.
- Mechanisms underlying the initiation and maintenance of AF vary greatly among cardiovascular diseases; autonomic denervation therefore can be beneficial or harmful.
- Autonomic denervation for each cardiovascular disease should only be performed with evidence from randomized clinical trials to demonstrate its efficacy and safety.

## INTRODUCTION

Postoperative atrial fibrillation (POAF) is one of the most common complications of cardiac surgery and is a multifactorial disease.^1,2^ Contributing factors include sympathetic surge, intense inflammation, oxidative stress and preexisting atrial diseases. Despite the use of β-blockers and amiodarone, the incidence of POAF remains high.^3-5^

Ganglionated plexi (GP), embedded in epicardial fat pads, are widely considered as the “integration centers” of the extensive neural network formed by the intrinsic cardiac autonomic nervous system (ANS).^6-9^ GP contain both sympathetic and parasympathetic neural elements. Preclinical and clinical studies demonstrated that simultaneous or sequential activation of cardiac parasympathetic and sympathetic systems preceded AF initiation.^10-12^ The four major atrial GP adjacent to pulmonary veins (PV) are particularly important in AF initiation and maintenance.^13,14^ Circumferential PV isolation transects at least 3 of the 4 major atrial GP. Autonomic denervation therefore has been proposed to play an important role in the success of AF ablation.^15,16^

It is known that excessive intracellular Ca^2+^ concentration can be lethal to neurons.^17,18^ In the CAP-AF (<u>C</u>alcium-mediated <u>A</u>utonomic Denervation in <u>P</u>ostoperative <u>A</u>trial <u>F</u>ibrillation) trial, we demonstrated that calcium-mediated neurotoxicity by injecting calcium chloride (CaCl_2_) into the four major atrial GP reduced the incidence of POAF by 63% as compared to the sham injection sodium chloride (NaCl) in patients undergoing coronary artery bypass grafting surgery (CABG).^19^ We hypothesized that the substrate of POAF and the role of ANS may be distinct among patients with different cardiovascular diseases. The CAP-AF2 trial was designed to investigate if Ca-mediated autonomic denervation could prevent POAF in patients with mitral regurgitation (MR) without persistent AF who underwent mitral valve (MV) replacement or repair.

## METHODS

### Trial Design and oversight

The <u>C</u>alcium-mediated <u>A</u>utonomic Denervation in <u>P</u>ostoperative <u>A</u>trial <u>F</u>ibrillation-2 (CAP-AF2) trial is a single-center, sham-controlled, double-blind, randomized clinical trial that was conducted in accordance with the principles of the Declaration of Helsinki and the International Conference on Harmonization Good Clinical Practice Guideline. The protocol was approved by the Ethics Committee of the General Hospital of Northern Theater Command and registered in the Chinese Clinical Trial Registry (registration number: ChiCTR2000029314; website: http://www.chictr.org.cn/showproj.aspx?proj=48587). The trial was overseen by the Ethics Committee and the Science Discipline Department. Both committees had the authority to stop the trial for safety or efficacy concerns. All patients enrolled into this trial gave informed consent for participation.

The primary outcome is the incidence of POAF in the first 7 days after surgery.^19^ POAF was defined as any atrial tachyarrhythmia episode^5^ lasting ≥30 seconds following surgery.^19^ Secondary outcomes include length of hospitalization following surgery, AF burden during the first 7 postoperative days, average ventricular rate during AF, percentage of patients with AF lasting ≥ 24 or 48 hours, actionable antiarrhythmic therapy to treat POAF, frequency of premature atrial contractions (PACs) as well as plasma level of inflammatory markers including interleukin-6 (IL-6) and high sensitivity c-reactive protein (hsCRP).

### Patient Population

Adult patients referred for isolated MV repair or replacement for MR were eligible for enrollment. Exclusion criteria included: (1) adult patients >80 years of age, (2) history of persistent AF or persistent AF detected by 7-day Holter monitoring before surgery, (3) prior cardiac surgery, 4) poor compliance or inability to complete follow-up, (5) urgent cardiac surgery, (6) congenital heart disease, (7) significant mitral stenosis (MV orifice area <2 cm^2^), (8) concomitant surgery of any kind, (9) abnormal liver or kidney function >3x upper normal limit, (10) diseases requiring radiotherapy, chemotherapy or long-term hormone treatment, (11) ejection fraction <40%, (12) taking class I or III antiarrhythmic agents before surgery, (13) significant left atrial enlargement (>5 cm), (14) concomitant AF ablation, (15) participating in another clinical trial and (16) refusal to enrollment.

### Sample Size Calculation and Randomization

We assumed a 40% incidence of POAF in the NaCl group based on the historical POAF incidence in the study center. We determined the enrollment of 152 CaCl_2_ patients and 152 NaCl patients would provide a power of at least 80% to detect a 15% absolute reduction in the incidence of POAF in the CaCl_2_ group compared to the NaCl (sham) group with a two-sided type-I error rate of 0.05. We expected a 5% drop-out rate; each group will therefore require 160 patients.

Eligible patients were randomly assigned to either the CaCl_2_ group or NaCl group by means of a computer-generated randomization system with a block of 8 with the group assignment concealed. CaCl_2_ or NaCl solution was prepared in a syringe accordingly. The surgery staff as well as the personnel involving post-operative care, clinical data collection and outcome assessment were blinded to randomization.

### Preoperative baseline data collection

Prior to surgery, all patients with severe MR referred for MV surgery received 7-day Holter monitoring with the capability of heart rate variability calculation.^19^ Patients with a history of persistent AF or had AF for ≥7 days preoperatively were excluded from this study. The rest of the patients meeting the inclusion criteria were considered potential candidates.

### Intervention

After MV repair or replacement, operators performed multi-point injection of 2 mL of 5% CaCl_2_ or 0.9% NaCl into the four major atrial GP as previously described.^19^ All the surgeons were already familiar with the location of the four major atrial GP from the CAP-AF trial. The anterior right GP is located near the annular aspect of the right superior PV-atrial junction; the inferior right GP at the inferior-posterior LA, below the right inferior PV.^15,19^ The superior left GP is situated near the junction of the left superior PV, left atrium (LA) and left pulmonary artery; the inferior left GP at the inferior-posterior LA below the left inferior PV.

### Outcome Measures

Continuous telemetry monitoring and 7-day Holter monitoring ensued after surgery. Continuous heart rate variability (HRV) was measured for 7 days following surgery. If AF ≥30 seconds was detected by any of the three modalities, it was considered a POAF event: (1) routine 12-lead ECG, (2) continuous telemetry monitoring with full-disclosure until discharge and (3) 7-day Holter monitoring equipped with an automated algorithm for detection of AF onset and duration as well as PACs and nonsustained atrial tachycardia as previously described (Yueguang Medical Technologies, Shanghai, China).^19^ POAF events were adjudicated by physicians who were blinded to randomization. AF burden was defined as the sum of the duration of all episodes of AF during the 7-day period after surgery. The standard practice to treat POAF in the study center was to initiate amiodarone therapy after AF sustained ≥ 30 minutes or AF caused hemodynamic concerns. In the presence of myocardial ischemia or hemodynamic compromise, POAF was treated with cardioversion or intravenous esmolol infusion, in addition to amiodarone therapy. Inflammatory markers including hsCRP and IL-6 were measured on postoperative day 1 and 3.

### Surgical specimen acquisition and immunohistochemistry

After midterm analysis, to better understand the mechanisms underlying the contradictory effect of Ca^2+^-mediated autonomic denervation, 113 consecutive patients referred for CABG or MV surgery were screened. Forty patients meet the inclusion criteria of the CAPF-AF or CAP-AF2 trial (CABG, n=20; MV surgery, n=20) gave consent to allow investigators to take tissue samples (average 0.5×0.5 cm^2^) in the anterior right GP and adjacent myocardium (**Online Table**). The samples were divided into two parts. One part of the samples was fixed in 4% paraformaldehyde for 4 hours and embedded in paraffin; sections were performed longitudinally and cut every 5 µm. The other part was frozen at -80°C for subsequent molecular biological assays.

### Immunohistochemistry studies

Immunohistochemistry of tyrosine hydroxylase (TH) and choline acetyltransferase (ChAT) was performed as previously describe.^20^ Image analysis of nerve density and immunohistochemistry staining was detected via ImageJ (Rawak Software Inc., Stuttgart, Germany). Quantitative analysis of the TH and ChAT expression in myocardial and ARGP samples was determined by ELISA and Western blot as described before.^20^

### Statistical Analysis

All analyses were based on the intention-to-treat principle including all patients who were randomized. For the primary outcome, Kaplan-Meier survival curves were used to describe the time-dependent incidence of POAF of both groups, the difference of which was compared by the log-rank test. Secondary outcomes were evaluated by Mann-Whitney U test, Chi-square test or Fisher’s exact test. Data were reported as medians and quartiles (1st, 3rd quartile) for quantitative data and as numbers for qualitative data. For the heart rate variability (HRV) data requiring repeated measures, generalized estimating equations were used to test possible differences in HRV, assuming treatment and time since surgery as fixed factors, with marginal distribution and a first-order autoregressive correlation matrix to test the main and interaction effects. All analyses were conducted using the SPSS software (version 24.0) and R software version 4.1.3. All reported *P* values are two-sided; significant level is 0.05. *P* values and 95% confidence intervals presented in this report have not been adjusted for multiplicity; therefore, inferences drawn from these statistics should be interpreted with caution.

## RESULTS

Between November 1, 2020 and March 29, 2022, 884 patients were referred to the General Hospital of Northern Theater Command for surgery to treat severe MR. A total of 160 patients, who met the inclusion criteria and gave consents, were randomized to the CaCl_2_ group (N=78) and NaCl group (N=82) before the midterm analysis (**Figure 1A**). Baseline characteristics were not different between the two groups (**Table 1**). Three of 78 patients in the CaCl_2_ group and 2 of 82 patients in the NaCl group had paroxysmal AF based on history and 7-day Holter monitoring before surgery.

**Figure 1.**
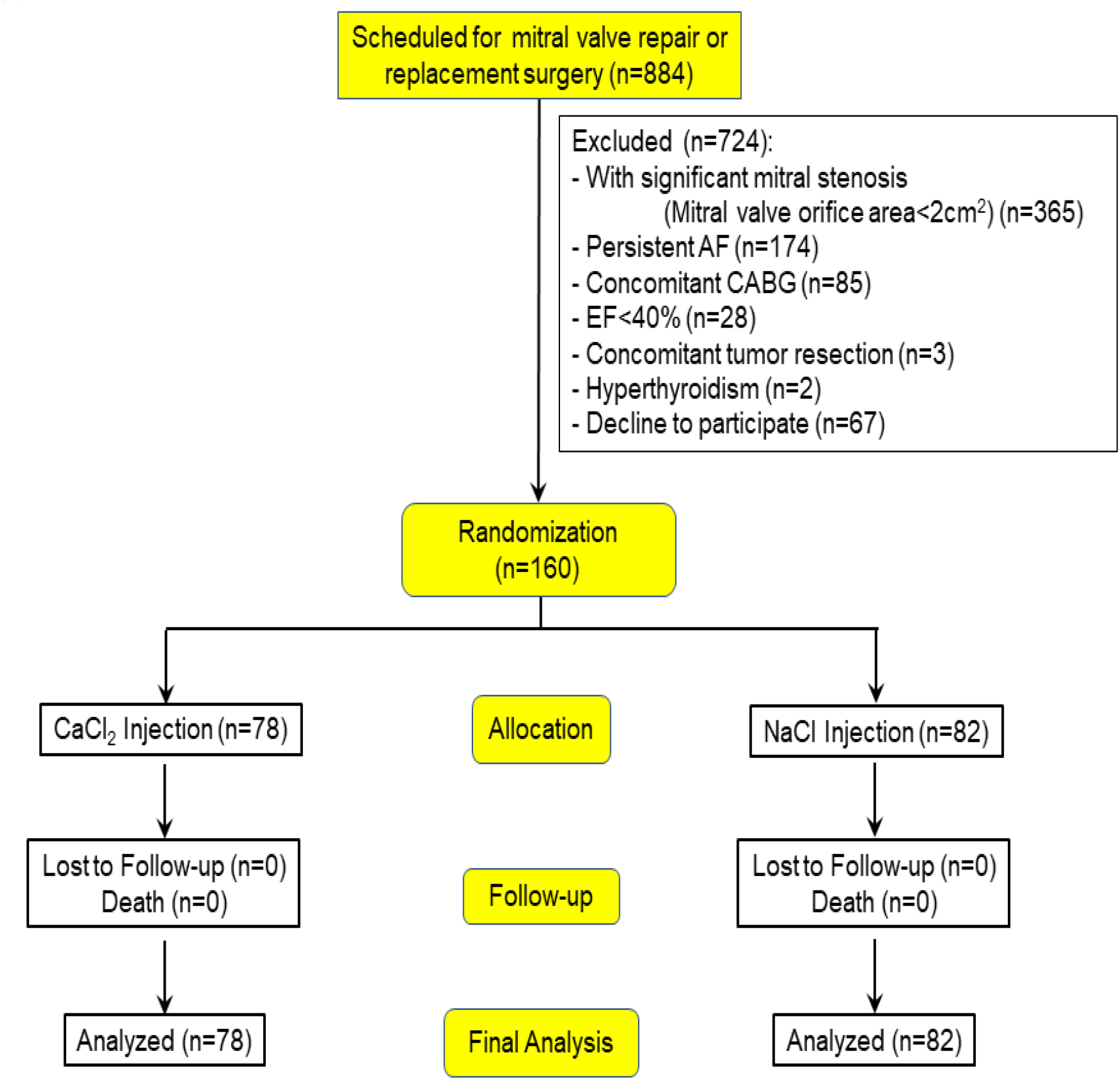
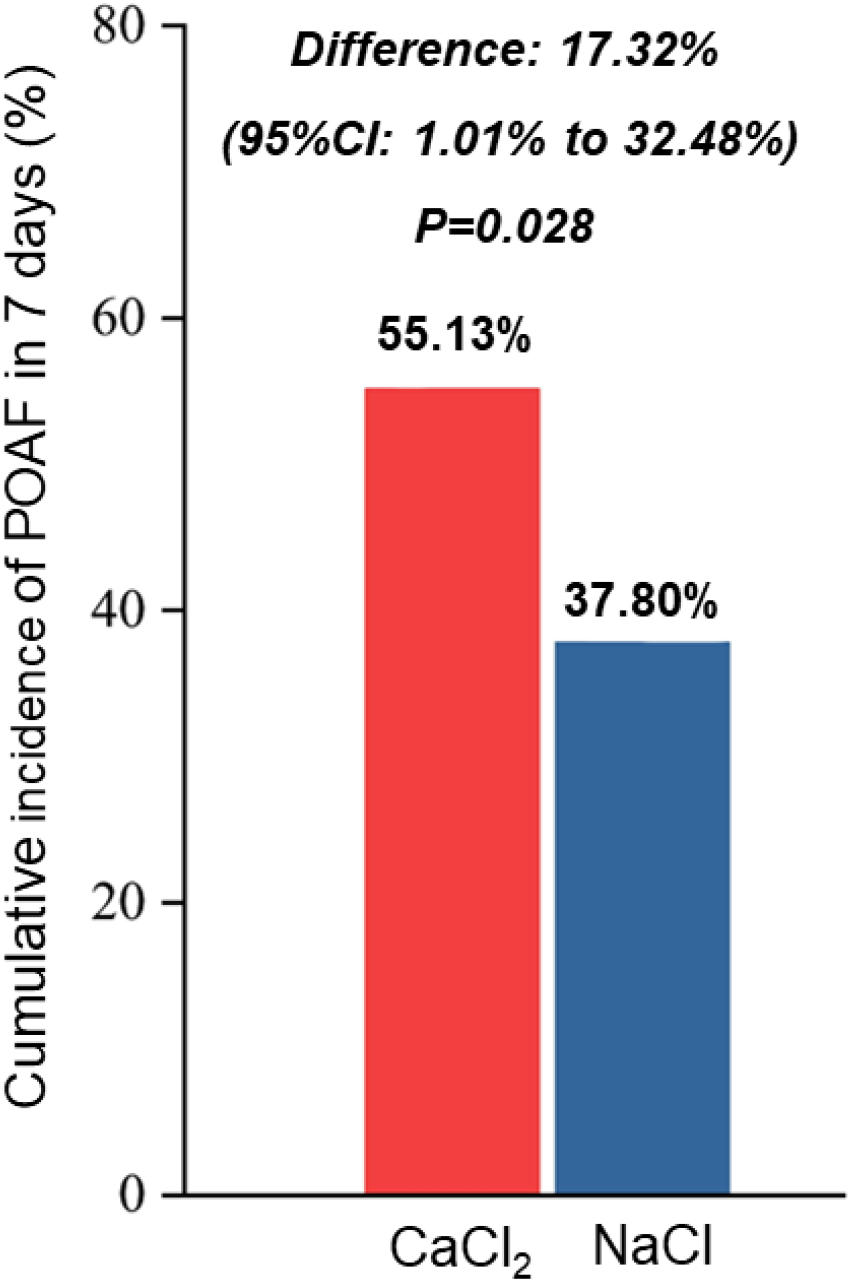
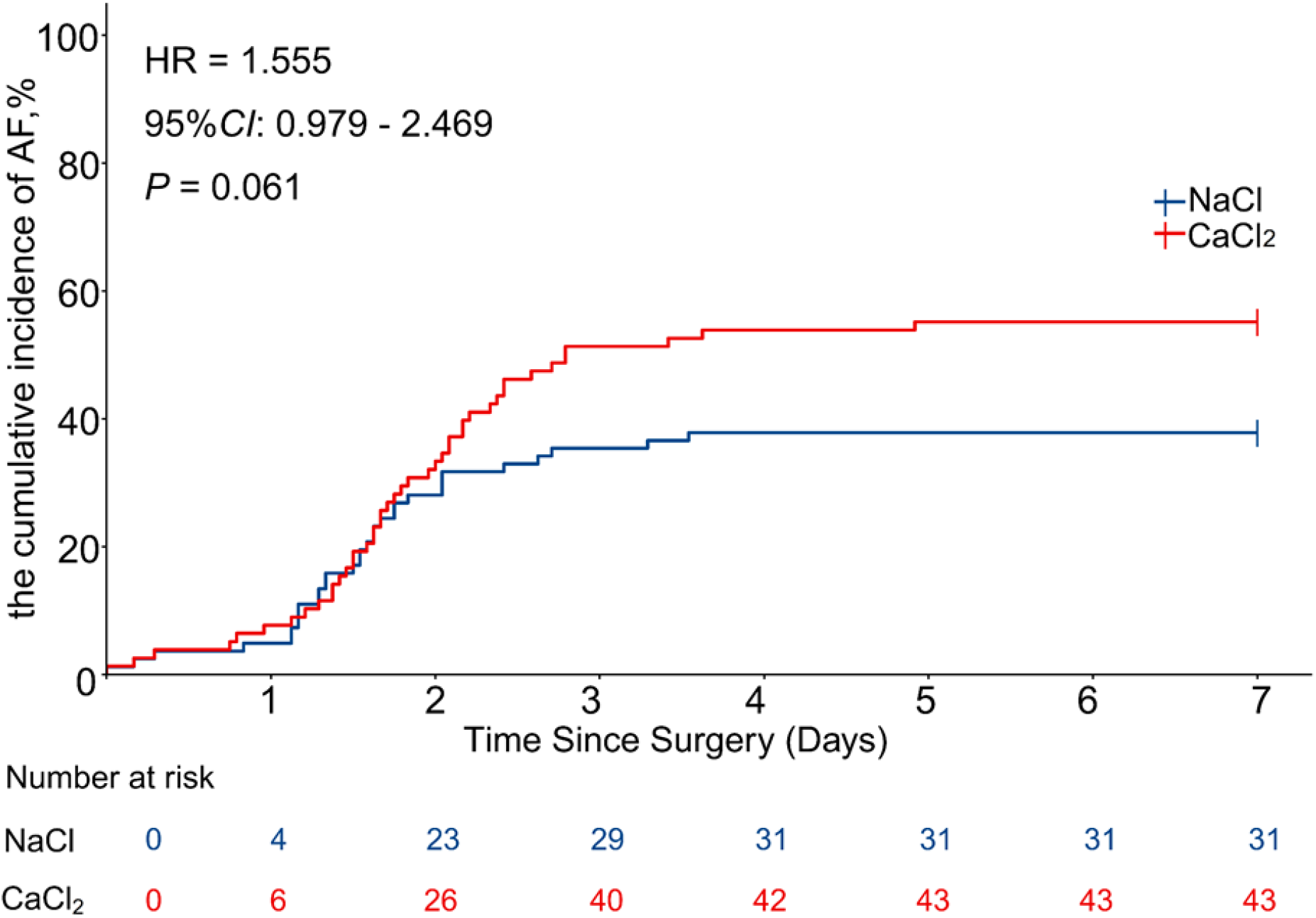
Trial design and primary outcome. A. Flow diagram of trial design. A total of 884 patients were screened. Up to the midterm analysis, 160 patients consented to participate. Participants were 1:1 randomized to CaCl_2_ injection and NaCl injection. **B.** Cumulative incidence of POAF over 7 days after surgery. **C.** Kaplan-Meier curve of POAF. HR: hazard ratio. CI: 95% confidence interval.

**Table 1.** Demographic and Clinical Characteristics at Baseline.

|  | **CaCl<sub>2</sub> (n=78)** | **NaCl (n=82)** | **P** |
| --- | --- | --- | --- |
| Age, yrs | 60 (56, 63) | 61 (53.25, 66) | 0.652 |
| Female, n (%) * | 32 (41.03) | 38 (46.34) | 0.498 |
| Hypertension, n (%) * | 33 (42.31) | 25 (30.49) | 0.120 |
| Diabetes, n (%) * | 9 (11.54) | 5 (6.1) | 0.223 |
| paroxysmal AF, n (%) * | 3 (3.85) | 2 (2.44) | 0.955 |
| CHA<sub>2</sub>DS<sub>2</sub>-VASc score | 1 (0,2) | 1 (1,2) | 0.235 |
| CAD, n (%) * | 18 (23.08) | 12 (14.63) | 0.171 |
| EF, % | 57 (55, 60) | 58 (56, 60) | 0.167 |
| FS, % | 30 (28, 32) | 30 (29, 32) | 0.093 |
| Left atrium, mm | 44 (40, 48) | 42 (38, 46.25) | 0.084 |
| On β-blocker, n (%) | 31 (39.7%) | 28 (34.2%) | 0.463 |
| Average HR, beats/min | 74 (69.5, 80) | 75 (70, 82) | 0.323 |
| Highest HR, beats/min | 117 (109, 127) | 118 (110, 130) | 0.227 |
| Lowest HR, beats/min | 51 (48, 57) | 54 (49, 59) | 0.069 |
| hsCRP, mg/l | 0.25 (0.25, 3.1) | 0.61 (0.25, 1.75) | 0.788 |
| IL6, pg/l | 3.08 (1.87, 6.68) | 2.73 (1.85, 4.88) | 0.838 |
| BNP, pg/l | 461.6 (107.9, 1622) | 223.2 (107.7, 764.32) | 0.850 |
| ALT, U/l | 19.7 (14.32, 26.65) | 17.2 (13.51, 27.56) | 0.390 |
| AST, U/l | 19.76 (15.35, 26.24) | 19.04 (16.74, 26.51) | 0.738 |
| Creatinine, mmol/l | 74.11 (62.61, 83.5) | 67.98 (60.83, 81.46) | 0.348 |
| Heart rate variability |  |  |  |
| pNN50, % | 3.35(1.42, 9) | 3.25(0.9, 9.75) | 0.391 |
| rMSSD, ms | 19.5(15.83, 29.4) | 19.85(13.83, 31.1) | 0.941 |
| SDNN, ms | 55.2 (45.7, 64.15) | 55.3 (42.1, 66.22) | 0.927 |
| Premature atrial beats |  |  |  |
| Single, % | 0.07 (0.02, 0.27) | 0.1 (0.02, 0.27) | 0.819 |
| Couplets, 10<sup>-2</sup>.% | 0.23 (0.04, 0.87) | 0.22 (0.05, 0.86) | 0.988 |
| NSAT, 10<sup>-2</sup>.% | 0.11 (0.03, 0.47) | 0.19 (0.03, 0.62) | 0.380 |

Values are median (1st, 3rd quartile), or n (percentage). AF= atrial fibrillation; ALT = alanine transaminase; AST = aspartate transaminase; BNP = brain natriuretic peptide; CAD: coronary artery disease; EF = ejection fraction; FS = Fraction shortening; COPD = chronic obstructive pulmonary disease; hsCRP = high-sensitivity C-reaction protein; IL-6 = interleukin-6; MI = history of myocardial infarction; pNN50 =percentage of RR intervals that differ by >50 ms; rMSSD =root-mean-square of successive RR interval difference; SDNN = SD of normal-to-normal interval; NSAT=non-sustained atrial tachyarrhythmia. \*: Calculated by chi-square test. Others: calculated by Mann-Whitney *U* test.

No patient received other pharmacological prophylaxis for POAF. All patients underwent successful surgeries for MV replacement or repair as previously described.^21^ Thirty-six and 45 patients underwent MV repair in the CaCl_2_ group and NaCl group, respectively. The rest of the patients received MV replacement. Under direct vision, 2 mL of CaCl_2_ or NaCl was injected into each of the 4 major atrial GP in all 160 patients as described previously.^19^ Injection of 4 GP took approximately 10 minutes. CaCl_2_ solution leak was quickly removed by suction to prevent possible injury to the surrounding tissue. After injection, the pericardial space was irrigated with saline before closure.

### Primary Outcome

Midterm analysis revealed that the POAF incidence was higher in the CaCl_2_ group (55.13%) than that in the NaCl group (37.80%; see results below). The research oversight committee determined that CaCl_2_ injection caused harm and terminated the trial. All the analyses therefore only included the 160 patients enrolled prior to the midterm analysis (78 and 82 patients in the CaCl_2_ and NaCl group, respectively). All 160 patients stayed in the hospital for ≥7 days; there was no missing data. POAF occurred during the first 7 days after surgery in 31 of 82 patients (37.80%) in the NaCl group and 43 of 78 patients (55.13%) in the CaCl_2_ group (95% confidence interval of difference: 1.01%-32.48%, *P*= 0.028) (**Figure 1B**). The Kaplan-Meier curve also illustrates separation of the two curves on postoperative day 3 when the occurrence of POAF typically reaches its peak (**Figure 1C**). The median of the first episode of POAF occurred 37.3 (28.1, 48.9) hours after surgery in the NaCl group and 41.5 hours (33.0, 56.9) in the CaCl_2_ group (*P*=0.132). All patients with AF received subcutaneous enoxaparin injection until discharge.

### Secondary Outcomes and AF-related measurements

The median length of hospitalization and ICU stay were not different between the two groups (**Table 2**). When POAF occurred, more patients in the CaCl_2_ group were hospitalized >7 days after surgery (69.8% vs. 45.2%, *P*=0.033). Patients in the CaCl_2_ group had more episodes of POAF (p=0.007) but with similar AF burden. The average ventricular rate in POAF as well as the number of patients with POAF>24 hours or >48 hours were not different between the two groups, either. More patients in the CaCl_2_ group required amiodarone to treat POAF, which was determined by clinicians when POAF lasted longer than 30 minutes. Plasma levels of hs-CRP and IL-6 on postoperative day 1 and 3 were not different between the two groups. Two patients in each group required cardioversion for hemodynamic compromise. The time-domain measurements as well as the frequency domain measurements of HRV such as low frequency (LF) power, high frequency (HF) power and the LF/HF ratio were not different between the two groups (**Online Figure 1**). The frequency of PACs, couplets and nonsustained atrial tachycardia was not different between the two groups on each postoperative day (**Online Figure 2**) as well.

**Table 2.** Secondary outcomes and peri-operative conditions.

|  | CaCl<sub>2</sub> (n=78) | NaCl (n=82) | p |
| --- | --- | --- | --- |
| ICU stay, hours | 24 (21, 37.25) | 23 (20, 37.75) | 0.799 |
| Hospitalization, days | 8 (7, 9) | 8 (7, 9) | 0.317 |
| Hospitalization > 7 days, n (%) * | 30 (69.8) | 14 (45.2) | **0.033** |
| AF burden, % | 2.01 (0, 25.07) | 0 (0, 10.24) | 0.066 |
| Number of AF episodes in patient | 1 (0, 4) | 0 (0, 1) | **0.007** |
| Average ventricular rate in AF | 103 (87, 127) | 108 (94, 125) | 0.349 |
| Patients with AF >48h, n (%) * | 11 (14.10) | 10 (12.20) | 0.721 |
| Patients with AF >24h, n (%) * | 19 (24.36) | 12 (14.63) | 0.120 |
| Inflammatory markers |  |  |  |
| hs-CRP (POD1), mg/L | 41.78 (16.83, 60.02) | 37.16 (23.57, 68.7) | 0.444 |
| IL6 (POD1), pg/L | 89.60 (63.90, 131.83) | 95.25 (58.62, 130.35) | 0.661 |
| Hs-CRP (POD3), mg/L | 119.47 (83.04, 169.62) | 122.62 (82.51, 178.97) | 0.500 |
| IL6 (POD3), pg/L | 62.94 (41.39, 99.46) | 71.72 (39.13, 103.67) | 0.965 |
| Amiodarone, n (%) * |  |  | **0.039** |
| 0 | 63 (80.77) | 75 (91.46) |  |
| 0-2500 mg | 10 (12.82) | 7 (8.54) |  |
| >2500 mg | 5 (6.41) | 0 (0) |  |
| Esmolol, n (%) * |  |  | 0.873 |
| 0 | 53 (67.95) | 55 (67.07) |  |
| 0-600 mg | 5 (6.41) | 7 (8.54) |  |
| >600 mg | 20 (25.64) | 20 (24.39) |  |
| Cardioversion, n (%) † | 2 (2.56) | 2 (2.44) | >0.999 |
| Major surgery time, min | 111 (90, 137) | 116 (91, 145) | 0.565 |
| Surgery type, n (%) * |  |  |  |
| MV repair | 36 (46.16) | 45 (54.88) | 0.270 |
| MV replacement | 42 (53.85) | 37 (45.12) |  |
| MV replacement, n (%) * |  |  |  |
| Mechanical valve | 28 (66.67) | 26 (70.27) | 0.731 |
| Tissue valve | 14 (33.33) | 11 (29.73) |  |
| Death, n (%) | 0 | 0 | - |
| Post-operative EF, % | 56 (53, 58) | 57 (55, 59) | 0.222 |
| Troponin-T, ng/L | 380(260.25, 559.75) | 333.5(243.75, 490.25) | 0.131 |
| Poor wound healing, n (%) | 0 | 0 | - |
| Procalcitonin (POD3), ng/mL | 0.14(0.11, 0.17) | 0.13(0.10, 0.15) | 0.083 |
| Pleural effusion, n (%) † | 1 (1.28) | 4 (4.88) | 0.368 |
| On ventilator >96 h, n (%) † | 1 (1.28) | 0 | 0.487 |
| Average heart rate (bpm) | 85 (79, 91) | 85 (76, 93) | 0.768 |
| Renal dysfunction, n (%) † | 1 (1.28) | 0 | 0.487 |
| Post-operative myocardial infarction, % | 0 | 0 | - |
| Stroke, % | 0 | 0 | - |

Values are median (1st, 3rd quartile) or n (percentage). POD: postoperative day. All abbreviations identical to Table 1. \*Calculated by chi-square test. †Calculated by Fisher exact test. Others: calculated by Mann-Whitney *U* test.

### Adverse effects and complications

To prevent adverse effects such as significant bradycardia (heart rate <40 bpm) as described in the CAP-AF trial, CaCl_2_ and NaCl were slowly injected into each GP to avoid pressure buildup inside the GP. No significant bradycardia (<40 bpm) was noticed in any patient. Surgery duration (beginning of sternotomy to completion of wound closure) and peri-operative mortality and morbidity were not different between the two groups (**Table 2**).

### Comparison of POAF initiation between the CAP-AF and CAP-AF2 trial

To understand the mechanism(s) responsible for the contradictory effect of Ca^2+^- mediated autonomic denervation on POAF, we compared the pattern of POAF initiation that was adjudicated as POAF in the CAP-AF and CAP-AF2 trials. In the CAP-AF2 trial, POAF initiation was immediately preceded by frequent atrial bigeminy, couplets or nonsustained AT (NSAT, ≥3 beats) in 33/43 (76.7%; **Figure 2A**) and 9/31 patients (29.0%) in the CaCl_2_ and NaCl group, respectively (*P*<0.001). The most common type of POAF initiation in the NaCl group was sinus rhythm with a single PAC (16/31 patients, 51.6%; **Figure 2B**), which only occurred in 6/43 patients (14.0%) in the CaCl_2_ group (*P*<0.001). On the contrary, in the CAP-AF trial, POAF initiation was preceded by frequent atrial bigeminy, couplets or NSAT in 11/15 patients (73.3%) and 22/36 patients (61.1%) in the CaCl_2_ and NaCl group, respectively (*P*=0.405). The second most common initiation pattern was sinus rhythm with a single PAC that occurred in 3/15 (20.0%) and 12/36 (33.3%) patients in the CaCl_2_ and NaCl group, respectively (*P*=0.341).

**Figure 2.**
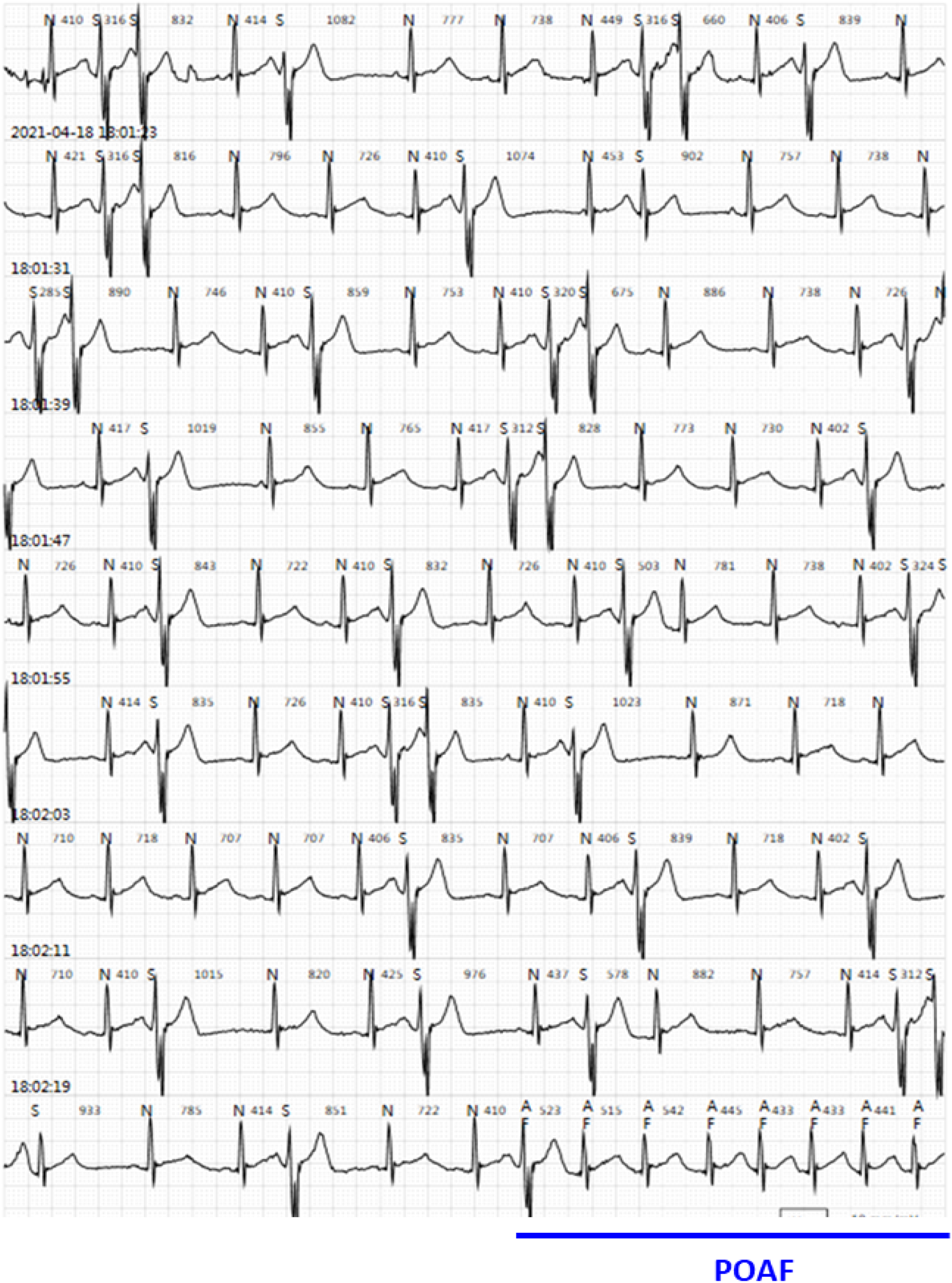
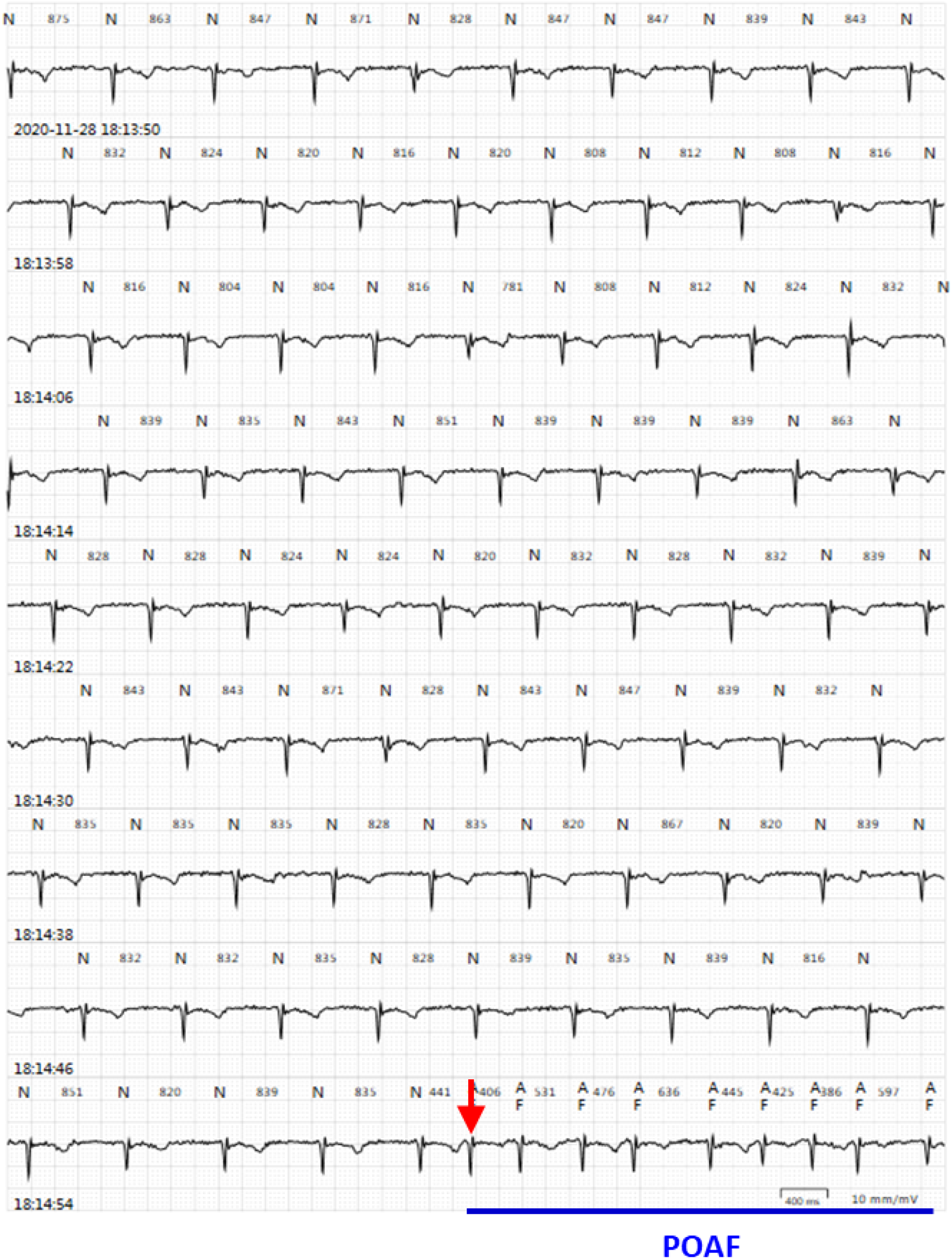
The most common pattern of POAF initiation in the CAP-AF2 trial. **A.** POAF was preceded by frequent atrial bigeminy, couplets or nonsustained AT in the CaCl_2_ group **B.** A single PAC (red arrow) initiated POAF in the NaCl group.

### Immunohistochemical staining of anterior right GP and adjacent atrial myocardium

Parasympathetic and sympathetic neural elements were marked by ChAT and TH staining, respectively. In the 40 patients not in the CAP-AF or CAP-AF2 trial (n=20 for CABG and n=20 MR surgery), the percentage of area in the anterior right GP stained positive for TH was significantly higher in the CABG group but that stained positive for ChAT was higher in the MV surgery group (**Figure 3A-C**). The ratio of TH/ChAT was therefore significantly higher in the CABG group. When quantitative analysis of TH and ChAT expressing nerves in the atrial myocardium adjacent to the ARGP was performed, similar results also hold true for nerve density (**Figure 3D-F)** as well as protein expression assessed by ELISA (**Figure 3G-I**) and Western blot (**Online Figure 3**). The extent of fibrosis in the atrial myocardium adjacent to the anterior right GP was not different between the two groups **(Online Figure 4**).

**Figure 3.**
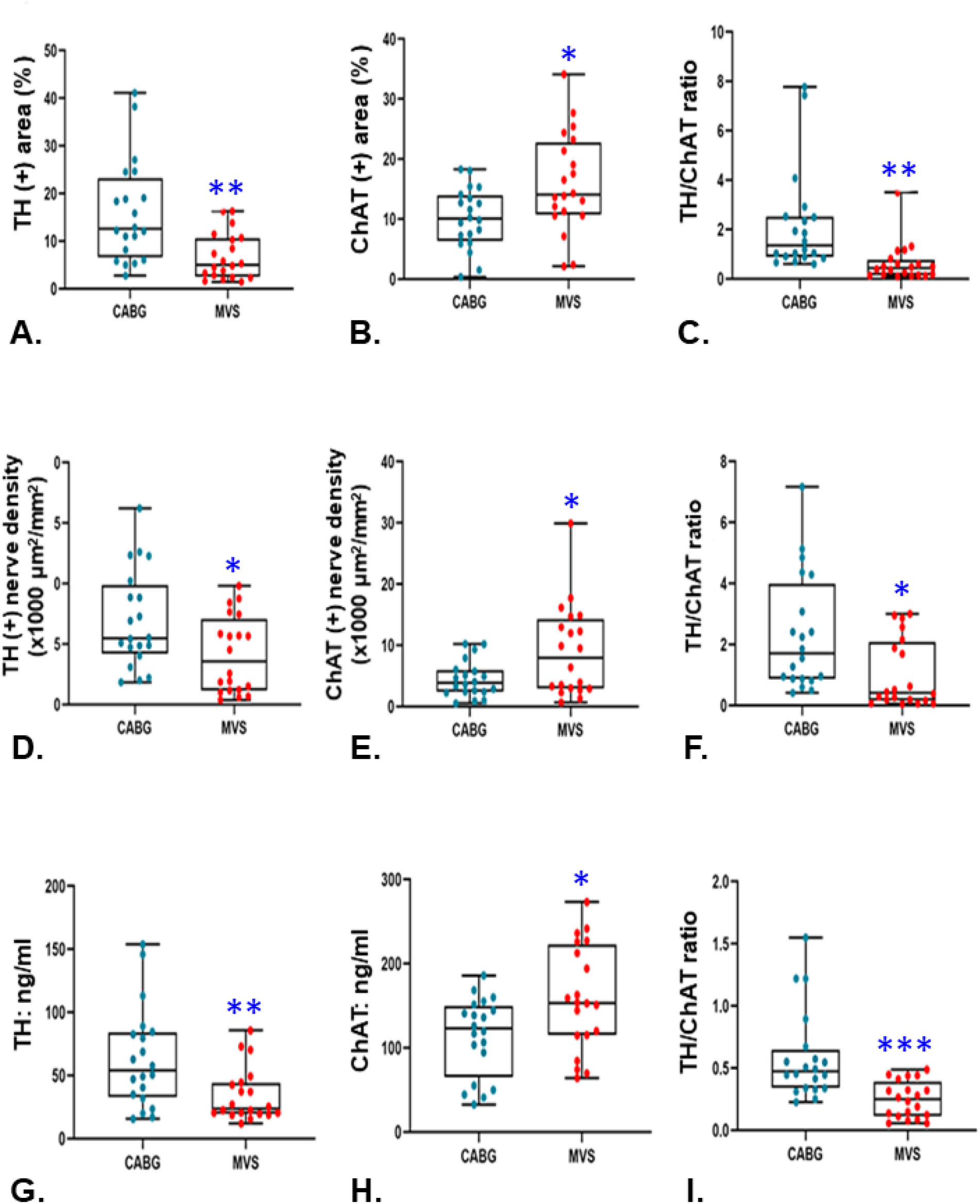
Tissue autonomic innervation. Tissue samples obtained from patients undergoing coronary artery bypass surgery (CABG, n=20) or mitral valve surgery (MVS, n=20) for mitral regurgitation. **A-C.** Percentage of the area in the anterior right ganglionated plexi stained positive for choline acetyltransferase (ChAT; **A**) or tyrosine hydroxylase (TH, **B**) as well as the TH/ChAT ratio (**C**). **D-F.** ChAT staining, TH staining and TH/ChAT ratio of autonomic nerves. **G-I.** Quantitative analysis by ELISA of TH and ChAT expression. **D-I.** Tissue samples obtained from atrial myocardium adjacent to the anterior right ganglionated plexi. GP. *: *p*<0.05; **: *p*<0.01; ***: *p*<0.001.

## DISCUSSION

The main finding of the CAP-AF2 trial is that Ca^2+^-mediated autonomic denervation unexpectedly increased the incidence of POAF in patients with MR without persistent AF, contradictory to the results of the CAP-AF trial that only recruited patients receiving isolated CABG. In the CAP-AF2 trial, atrial bigeminy, couplets or NSAT immediately preceded the initiation of POAF in the CaCl_2_ group but not the NaCl group. Immunohistochemistry studies demonstrated different autonomic substrates between patients with MR and ischemic heart disease. A lower ratio of TH/ChAT in the anterior right GP and adjacent myocardium in the former suggests parasympathetic dominance at the tissue level; a higher TH/ChAT ratio in the latter suggests sympathetic dominance. Ca^2+^-mediated autonomic denervation therefore exerts contradictory effects on the incidence of POAF on patients receiving MV surgery vs. CABG.

Multiple preclinical studies of MR-related AF have demonstrated similar findings.^22-27^ First, the atrial effective period was prolonged with significant heterogeneity. Second, moderate but heterogeneous interstitial fibrosis existed. Third, atrial conduction velocity was reduced and often heterogeneous. Fourth, inflammatory cell infiltration of atrial myocardium was observed. Fifth, malfunction of major metabolic pathways occurred in patients with MR and was further augmented by β-adrenergic stimulation. Improvement of metabolic derangement mitigated AF. Although spontaneous AF was rarely reported by these studies, AF could easily be induced by electrical stimulation, indicating that in the presence of appropriate triggers, AF can be initiated and maintained by the aforementioned AF substrates. In the present study, patients with persistent AF were excluded. The study population therefore represents one in which MR was severe enough that already created a substrate for AF but without appropriate triggers to initiate AF. Parasympathetic dominance at the tissue level may be protective in patients with severe MR. Disturbance of the cardiac autonomic balance by Ca^++^-mediate autonomic denervation provided the triggers to initiate POAF as illustrated in **Figure 2A.**

In patients with paroxysmal AF without significant structural heart diseases, AF episodes are commonly preceded by co-activation of the sympathetic and parasympathetic nervous systems. ^10,12,28^ However, exaggerated sympathetic activity is the hallmark of AF initiation. In the majority of patients including those with structural heart diseases, AF episodes are usually preceded by an increased low-frequency component of HRV, a marker of sympathetic hyperactivity.^28^ Preclinical studies on ambulatory animals also indicated that paroxysmal AF and AT were invariably preceded by activation of the intrinsic cardiac ANS evidenced by rapid discharges recorded from the GP and ligament of Marshall.^11,29^ Although the paroxysmal AF or AT episodes were also accompanied by co-activation of the extrinsic cardiac ANS, such as vagus nerve and stellate ganglion, 20-25% of the episodes of paroxysmal AT or AF were not preceded by activation of the vagus nerve or stellate ganglion, indicating that the intrinsic cardiac ANS (e.g. GP) plays an indispensable role in the initiation of paroxysmal AF or AT. Moreover, PACs were typically preceded by a surge of sympathetic nerve activity; most of the paroxysmal AF or AT episodes followed a larger and more sustained surge of sympathetic nerve activity.^11,30^ The average activity of the intrinsic cardia ANS, evidenced by direct neural recording of GP, also strongly correlated with the PAC burden in patients after cardiac surgery.^30^

In the CAP-AF2 trial, POAF was preceded by frequent atrial bigeminy, couplets or NSAT in only 9/31 (29.0%) patients in the NaCl group, in contrast to 33/43 (76.7%) patients in the CaCl_2_ group. That is, CaCl_2_-mediated autonomic denervation altered the POAF initiation pattern from sinus rhythm with a single PAC to frequent bigeminy, couplets or NSAT, possibly resulting from shifting the sympathetic-parasympathetic balance (**Figure 2**). As discussed before, PACs and paroxysmal AT are often preceded by a surge of sympathetic nerve activity. Immunohistochemistry data revealed parasympathetic dominance at the tissue level (both GP and atrial myocardium) in patients receiving MV surgery. Ca^2+^-mediated autonomic denervation likely injured more parasympathetic than sympathetic neural elements, which tipped the balance toward sympathetic dominance. As a result, frequent atrial bigeminy, couplets or NSAT occurred, eventually initiating POAF. Of note, in the CAP-AF trial, Ca^2+^-mediated autonomic denervation suppressed PACs and NSAT but did not affect the initiation pattern of POAF. We hypothesize that a higher sympathetic state is also critical in POAF initiation in patients undergoing CABG. Ca^2+^-mediated autonomic denervation mitigated higher tissue sympathetic activity, evidence by less frequent complex PACs and NSAT. Additionally, HRV showed diminished HF and LF components but preserved LF/HF ratio,^19^ suggesting that the sympathetic-parasympathetic balance was not perturbed by autonomic denervation thereby not altering the initiation pattern of POAF. Therefore, for patients in the CaCl_2_ group of the CAP-AF trial, if autonomic denervation failed to overcome the higher sympathetic activity, POAF was triggered by similar mechanisms as seen in the NaCl group. Based on the contradictory results in the CAP-AF and CAP-AF2 trials, perturbation of the autonomic balance can be beneficial or harmful, depending on the underlying substrate.

An important lesson learned from the two CAP-AF trials is that autonomic remodeling and perhaps other remodeling processes may vary greatly among different cardiovascular diseases, leading to differential effects on POAF when cardiac ANS is perturbed. If the CAP-AF2 trial had recruited both MV surgery and CABG populations, the results might have been neural; the harm to the MV surgery population would have not been exposed. Future studies on the pathogenesis or therapies of AF, not limited to POAF, may need to focus on a relatively homogeneous population and avoid a study population including multiple cardiovascular diseases. For example, the AFACT^31^ trial randomized patients to surgical PVI vs. surgical PVI+GP ablation and reported no benefit conferred by GP ablation, contradictory to the beneficial effects of GP ablation reported by the largest randomized clinical trial (catheter PVI vs. PVI+GP ablation) that only recruited patients with paroxysmal AF without prior ablation.^16^ Notably, in the AFACT trial, 59% patients had persistent AF, 43% patients had severely enlarged left atrium (LA volume index>40 mL/M^2^), and nearly 25% had prior catheter ablation. The conflicting results from the aforementioned two trials may stem from two distinct study populations. Two recently published trials further exemplified this viewpoint. By injecting botulinum toxin into the major atrial GP as a means of autonomic denervation, reduction of the incidence of POAF was reported in one randomized clinical trial on patients receiving CABG but a neutral effect was reported by the other trial recruiting patients undergoing CABG and/or valve surgery.^32, 33^

### Study Limitation

First, we did not directly record the neural activity of the GP; the activity of the intrinsic cardiac ANS was therefore deduced from expression of TH and ChAT in the anterior right GP and adjacent myocardium. In addition, the postoperative HRV was inconclusive likely due to a smaller-than-expected sample size resulting from premature termination of the trial. The interpretation that higher sympathetic activity preceded POAF was therefore based on published pre-clinical and clinical studies. Second, we do not have immunohistochemical data on normal controls to compare tissue autonomic innervation among normal controls, patients with MR and patients with ischemic heart disease. Therefore, sympathetic and parasympathetic dominance described in this study only represents the comparison between the MR and CABG populations. Third, for logistical reasons, only the anterior right GP and adjacent myocardium were sampled. Tissue autonomic innervation and fibrosis may not be generalized to the entire atrial neural network and myocardium. Fourth, only five of 160 patients were found to have paroxysmal AF based on history and 7-day Holter monitoring prior to surgery. The prevalence of paroxysmal AF is probably underestimated. We presume that under-diagnosis of paroxysmal AF would have been evenly distributed by randomization and will not affect the outcome assessment.

## CONCLUSIONS

Unlike patients undergoing CABG, autonomic denervation increased the risk of POAF in patients receiving MV surgery, indicating distinct substrates for POAF in different cardiovascular diseases. This observation may explain conflicting results from published POAF trials that recruited patients undergoing CABG and valvular surgery. While autonomic denervation has emerged as a novel therapy to treat various cardiovascular diseases, it should only be performed with evidence supported by randomized clinical trials.

## Data Availability

After manuscript is accepted for publication, the de-identified data can be available to other researchers after an application approved by the corresponding authors

## Abbreviations

ANS: autonomic nervous system
CABG: coronary artery bypass grafting
CaCl_2_: calcium chloride
ChAT: choline acetyltransferase
GP: ganglionated plexi
MV: mitral valve
NaCl: sodium chloride
POAF: postoperative atrial fibrillation
TH: tyrosine hydroxylase

